# Short-Term Patient-Reported Outcomes After Facial Skin Cancer Surgery: A Prospective Longitudinal Study Using the FACE-Q Skin Cancer Module

**DOI:** 10.64898/2026.04.06.26349979

**Authors:** Maarten M.J. Ottenhof, Maarten M. Hoogbergen, René R.W.J. van der Hulst

## Abstract

**Background:** Patient-reported outcome measures provide essential data on treatment quality across diverse populations. The FACE-Q Skin Cancer Module was developed to assess outcomes specific to facial skin cancer patients. Longitudinal data characterizing outcome trajectories from surgery through early recovery remain limited.

**Objective:** We tracked how patient outcomes change from preoperatively through three months after surgery using the FACE-Q Skin Cancer Module in a prospective cohort of 288 patients undergoing facial skin cancer surgery.

**Methods:** Participants completed the module preoperatively and at 1 week and 3 months postoperatively. Five scales were evaluated: Appearance, Psychosocial Distress, Cancer Worry, Scars, and Adverse Effects. Friedman tests assessed overall change across timepoints; paired t-tests and Wilcoxon signed-rank tests evaluated pairwise comparisons.

**Results:** Of 288 enrolled patients (mean age 68.6±11.9 years, 46.5% female), 252 (87.5%) and 220 (76.4%) completed 1-week and 3-month follow-up, respectively. Facial appearance declined at 1 week (55.6 to 52.0, p=0.005) and returned to baseline by 3 months (57.0, p=0.274). Psychosocial distress increased acutely (14.5 to 19.0, p<0.001) with partial recovery at 3 months (17.1, p=0.012). Cancer worry decreased substantially (Δ=−7.8, SRM=−0.54, p<0.001), and scar satisfaction improved from 1 week to 3 months (Δ=+9.4, SRM=0.54, p<0.001). Adverse effects showed the largest improvement (Δ=−12.8, SRM=−0.88, p<0.001). Women showed less improvement in facial appearance than men (Δ=−2.2 vs +4.9, p=0.022). Clinical meaningfulness was assessed using minimally important difference thresholds: 36.9% of patients achieved meaningful improvement in appearance, 39.6% remained stable, and 23.4% experienced meaningful deterioration.

**Conclusions:** Short-term outcomes follow a predictable pattern, with acute perioperative worsening followed by recovery by 3 months for most patients.

## INTRODUCTION

Facial skin cancer represents the most common malignancy worldwide, with incidence rates increasing across most developed nations. Surgical excision remains the primary treatment approach, often necessitating reconstruction of the surgical defect. Although oncological outcomes are consistently favorable, recognition has grown that quality of life after surgery— encompassing facial appearance, psychosocial functioning, and cancer-related worry—merits equal clinical attention.

Patient-reported outcome measures capture the subjective experience of patients navigating disease and treatment. The FACE-Q Skin Cancer Module was developed specifically for this population, including independent scales that assess satisfaction with appearance, psychosocial distress, cancer worry, scar satisfaction, treatment-related adverse effects, and sun protection practices. Psychometric validation showed sound internal consistency, adequate construct validity, and responsiveness to clinical change.

Despite availability of this validated instrument, longitudinal data characterizing recovery patterns remain sparse. The typical questions persist: How do patients actually recover across the postoperative period? At what time points do patients experience maximal distress, and when does functional and psychological improvement emerge? Such information holds direct relevance for preoperative counseling and patient expectation management. Cross-sectional or single-timepoint assessments cannot capture the temporal arc of recovery that patients and clinicians need to understand.

We designed this study to characterize how patient-reported outcomes change from preoperatively through three months after facial skin cancer surgery using the FACE-Q Skin Cancer Module. Our specific objectives were to describe the overall pattern of change across five scales, to quantify the magnitude of change using standardized effect size metrics, to classify individual patient trajectories using minimally important difference thresholds, and to identify patient characteristics associated with differential recovery patterns.

## METHODS

### Study Design and Population

This prospective longitudinal cohort study enrolled consecutive patients scheduled for surgical treatment of facial skin cancer at a tertiary academic medical center. Inclusion criteria were age of 18 years or older, histologically confirmed facial skin malignancy, and scheduled surgical excision with or without reconstruction. Patients unable to complete questionnaires in Dutch were excluded from enrollment. The institutional review board approved the study protocol, and all participants provided written informed consent prior to enrollment.

### Outcome Measures

The FACE-Q Skin Cancer Module was administered at three designated timepoints: preoperatively (baseline), one week postoperatively, and three months postoperatively. Five scales were assessed: (1) Satisfaction with Facial Appearance (9 items, range 9–45), (2) Psychosocial Distress (8 items, range 8–40; higher scores indicate greater distress), (3) Cancer Worry (10 items, range 10–50; higher scores indicate greater worry), (4) Satisfaction with Scars (8 items, range 8–40; administered from week 1 onward), and (5) Adverse Effects of Treatment (10 items, range 10–40; administered from week 1 onward; higher scores indicate more effects). All raw scores were linearly transformed to a 0–100 scale for standardized interpretation using the formula: (raw sum − minimum) / (maximum − minimum) × 100.

### Statistical Analysis

Descriptive statistics were reported as means with standard deviations and medians with interquartile ranges. For assessment of change across three timepoints on continuous scales, Friedman tests were employed to avoid assumptions of normality. Pairwise comparisons between timepoints were conducted using paired t-tests and Wilcoxon signed-rank tests as appropriate. Effect sizes were calculated using Cohen’s d and the Standardized Response Mean (SRM), with the following classifications: small effects 0.20–0.49, medium effects 0.50–0.79, and large effects ≥0.80.

Individual patient trajectories were classified as improved, stable, or worsened on the basis of minimally important difference (MID) thresholds established from prior literature. For the Appearance scale, an MID of 5.6 points was applied. For Psychosocial Distress and Cancer Worry scales, an MID of 6.2 points was used.

Subgroup analyses were performed using Mann–Whitney U tests to compare outcomes by sex and Kruskal–Wallis tests for age groups (<60, 60–70, >70 years). Floor and ceiling effects were assessed at each timepoint. Baseline characteristics were compared between participants with complete three-month data and those lost to follow-up to evaluate potential attrition bias. All analyses were conducted using Python 3.11 with SciPy 1.11. Statistical significance was set at p<0.05 (two-sided). Multiple comparison corrections were not applied because subgroup analyses were exploratory in nature.

## RESULTS

### Study Population and Attrition

We enrolled 288 patients with a mean age of 68.6 years, of whom 46.5% were female and 27.8% reported at least one comorbidity. One-week follow-up was completed by 252 patients (87.5%), and three-month follow-up by 220 patients (76.4%). Non-completers were significantly older than completers (72.3 versus 67.4 years, p=0.003). Sex distribution did not differ between completers and non-completers (p=0.226), nor did the prevalence of comorbidities (p=1.000). Follow-up at one year was attempted via mailed questionnaire to 168 patients; only one survey was returned, yielding a response rate of 0.6% that precluded further long-term analysis.

*[Table 1. Patient Demographics]*

### Longitudinal Outcomes

#### Facial Appearance

Satisfaction with facial appearance showed a characteristic dip at one week followed by recovery by three months. Scores declined from 55.6 at baseline to 52.0 at one week (p=0.005). By three months, scores had recovered to 57.0, which was statistically equivalent to baseline (p=0.274 versus baseline). The improvement from one week to three months was significant (p=0.004). The Friedman test indicated significant overall change across the three timepoints (χ^2^=16.87, p<0.001).

#### Psychosocial Distress

Psychosocial distress increased acutely following surgery. Baseline scores of 14.5 rose to 19.0 at one week (p<0.001). At three months, distress scores declined to 17.1, though this remained elevated compared with baseline (p=0.012). The Friedman test confirmed significant change across timepoints (χ^2^=7.43, p=0.024).

#### Cancer Worry

Cancer worry decreased substantially from baseline to three months (25.2 to 17.2, p<0.001, SRM=−0.54), representing a medium effect. Surgical removal of the lesion with confirmed clear margins appeared to provide substantial psychological relief regarding cancer risk.

#### Scars and Adverse Effects

Satisfaction with scars showed marked improvement from one week to three months (53.6 to 62.0, p<0.001). Adverse effects showed the most substantial improvement among all scales, declining from 29.4 at one week to 16.0 at three months (p<0.001, SRM=−0.88), reflecting a large effect size. Resolution of postoperative symptoms such as swelling, bruising, sensory changes, and drainage accounted for this substantial improvement.

*[Table 2. FACE-Q Scores Across Timepoints (0–100 Scale)]*

Values are mean±SD (n). †Higher scores indicate worse outcomes. 𝕊Paired t-test (2 timepoints only). χ^2^ = Friedman test statistic.

*[Table 3. Effect Sizes for Pairwise Comparisons]*

†Higher scores indicate worse outcomes (negative change = improvement). SRM = Standardized Response Mean.

#### Individual Patient Trajectories

Individual patient trajectories from baseline to three months were classified using minimally important difference thresholds (Table 4). For facial appearance (MID=5.6), 41 patients (36.9%) showed clinically meaningful improvement, 44 patients (39.6%) remained stable, and 26 patients (23.4%) experienced meaningful deterioration. For psychosocial distress (MID=6.2), 57 patients (51.4%) remained stable, 32 patients (28.8%) reported meaningful worsening, and 22 patients (19.8%) experienced meaningful improvement. Cancer worry showed the most favorable distribution, with 58 patients (52.3%) achieving meaningful improvement, 36 patients (32.4%) remaining stable, and 17 patients (15.3%) reporting increased worry.

*[Table 4. Individual Patient Trajectories (Pre to 3 Months)]*

n=111 for all scales. †Higher scores = worse outcomes; for these scales, ‘improved’ = decrease > MID, ‘worsened’ = increase > MID. *For Psychosocial Distress, ‘improved’ means distress decreased and ‘worsened’ means distress increased.

#### Subgroup Analysis by Sex

Sex-stratified analysis revealed a significant difference in facial appearance outcomes. Men showed mean improvement of +4.9±15.1 points from baseline to three months (n=60), whereas women showed mean decline of −2.2±16.0 points (n=51; Mann–Whitney U=1916, p=0.022). No significant sex differences were observed for psychosocial distress (p=0.386) or cancer worry (p=0.819).

#### Subgroup Analysis by Age

Age-stratified analysis across groups (<60, 60–70, >70 years) revealed no statistically significant differences in change scores for any FACE-Q scale (all Kruskal–Wallis p>0.05). Although a trend toward greater cancer worry reduction appeared in the 60–70 age group (Δ=−12.7 points) compared with the >70 age group (Δ=−5.3 points), this difference did not achieve statistical significance (p=0.069).

#### Floor and Ceiling Effects

Floor and ceiling effects were minimal across scales. At baseline, floor effects were documented for Cancer Worry (6.2%) and Psychosocial Distress (2.3%), reflecting patients without baseline worry or distress. Floor effects increased at three months for Cancer Worry (20.4%) and Adverse Effects (15.9%), consistent with successful treatment outcomes. No ceiling effects were observed at any timepoint.

## DISCUSSION

Our investigation tracked how patient-reported outcomes evolved after facial skin cancer surgery using the FACE-Q Skin Cancer Module. Results revealed a consistent postoperative pattern: acute worsening at one week, followed by recovery toward or beyond baseline by three months. Yet considerable heterogeneity existed—both across outcome domains and among individual patients.

Facial appearance satisfaction declined at one week then recovered by three months. The trajectory makes intuitive sense: acute surgical edema and ecchymosis reduce appearance satisfaction early in recovery. Wound healing and resolution of swelling allow appearance satisfaction to return. Yet—and this matters—the group average obscures critical individual variation. Nearly one-quarter of patients (23.4%) experienced meaningful deterioration in appearance satisfaction that persisted through three months. For these patients, counseling focused solely on average outcomes proves misleading and fails to acknowledge realistic risk.

Psychosocial distress followed a similar arc—worse at one week, partially improved by three months—but never returned fully to baseline. The acute spike likely reflects multiple stressors: the physical wound itself, direct visualization of the surgical defect, and processing the cancer diagnosis anew. Incomplete resolution by three months suggests prolonged psychological adjustment is common; some patients may benefit from additional psychosocial support beyond the perioperative period.

Cancer worry improved most dramatically. Over half the patients experienced clinically meaningful reduction in cancer-related worry by three months. This outcome aligns with clinical intuition: once the tumor is removed and margins confirmed clear, immediate cancer fear diminishes substantially. Yet 15% of patients reported increased worry—a group likely preoccupied with recurrence risk or surveillance, both realistic concerns given baseline skin cancer recurrence rates.

Adverse effects and scar satisfaction improved markedly from one week to three months—the most substantial improvement among all measured scales. Postoperative symptoms (edema, ecchymosis, paresthesia, serous drainage) resolve gradually; patients perceive these improvements acutely. This trajectory informs realistic early recovery expectations.

We found a striking sex difference in facial appearance outcomes. Men improved an average of 4.9 points from baseline to three months; women declined 2.2 points. The mechanisms warrant consideration. Women may maintain higher appearance standards, experience greater societal pressure regarding facial aesthetics, or employ different coping strategies. Prior work documenting sex differences in appearance-related distress after facial procedures supports this observation. Surgeons should anticipate this differential trajectory and may need to provide targeted counseling or psychosocial support for female patients.

## Limitations

This investigation has several limitations. One-year follow-up collapsed: only one of 168 mailed questionnaires was returned (0.6% response rate), eliminating any assessment of sustained or delayed improvement. Attrition at three months was uneven—older patients dropped out at higher rates—which may bias three-month findings toward more optimistic outcomes in younger populations. We did not collect tumor characteristics (histology, size, depth), surgical approach, or reconstruction method, limiting exploration of procedure-specific subgroups. We did not apply correction for multiple comparisons in subgroup analyses, increasing type I error in those exploratory comparisons. Finally, data were collected at a single tertiary academic center, potentially limiting generalizability to other surgical settings or healthcare systems.

## Conclusions

Facial skin cancer surgery shows a predictable short-term recovery pattern. Patients experience worsened appearance satisfaction and increased psychosocial distress at one week—the consequence of surgical trauma. By three months, most patients recover; cancer worry and adverse effects improve substantially. Recovery isn’t uniform, though. Approximately one-quarter of patients don’t regain baseline appearance satisfaction. Women consistently show worse appearance outcomes than men. These findings can reshape preoperative counseling: surgeons now have evidence-based patterns to share with patients. Future work should extend follow-up beyond three months, examine specific surgical approaches and reconstruction types, and investigate mechanisms underlying sex differences in appearance satisfaction.

## Data Availability

The data underlying this study were collected at Clinique Rebelle, Amsterdam. Requests may be directed to the corresponding author. Clinic information: https://www.cliniquerebelle.com

## Notes

### Competing Interest Statement

The authors have declared no competing interest.

### Funding Statement

This study did not receive any funding.

### Author Declarations

The Medical Ethics Review Committee of the Amsterdam UMC, Amsterdam, The Netherlands, approved this study. All participants provided written informed consent.

