## supplemental tables for "Short-Term Patient-Reported Outcomes After Facial Skin Cancer Surgery: A Prospective Longitudinal Study Using the FACE-Q Skin Cancer Module"

MEDRXIV/2026/349979

**Table 1. Patient Characteristics (n = 146 with Complete Four-Timepoint Data)**

| **Characteristic** | **n (%)** |
| --- | --- |
| **Total patients with complete data** | 146 |
| **Sex** |  |
| Female | 74 (50.7%) |
| Male | 72 (49.3%) |
| **Age, years (mean ± SD)** | 67.5 ± 12.0 |
| **Surgical procedure** |  |
| Mohs micrographic surgery | 99 (67.8%) |
| Wide local excision | 35 (24.0%) |
| Combined approaches | 12 (8.2%) |
| **Lesion location** |  |
| Face and neck | 144 (98.6%) |
| Other | 2 (1.4%) |

*Values are n (%) unless otherwise specified. Age presented as mean ± standard deviation.*

**Table 2. Longitudinal FACE-Q Skin Cancer Module Scores Across Four Timepoints**

| **Scale / Timepoint** | **n** | **Mean ± SD** | **t-statistic** | **p-value** | **Cohen’s d** |
| --- | --- | --- | --- | --- | --- |
| **Appearance Concerns (range 9–36)** |  |  |  |  |  |
| Pre-operative | 146 | 29.9 ± 5.2 | — | — | — |
| 1 week post-op | 154 | 27.4 ± 6.4 | 4.82 | <0.001 | −0.52 |
| 3 months post-op | 146 | 30.1 ± 5.4 | −0.27 | 0.79 | 0.02 |
| 1 year post-op | 146 | 31.0 ± 5.5 | −2.10 | 0.037 | 0.19 |
| **Psychosocial Distress (range 8–32)** |  |  |  |  |  |
| Pre-operative | 146 | 12.3 ± 3.1 | — | — | — |
| 1 week post-op | 151 | 14.2 ± 5.2 | −4.40 | <0.001 | 0.55 |
| 3 months post-op | 146 | 12.9 ± 4.3 | −1.64 | 0.10 | 0.16 |
| 1 year post-op | 146 | 12.5 ± 3.5 | −0.65 | 0.52 | 0.05 |

*Paired t-tests comparing each follow-up timepoint to pre-operative baseline. d = Cohen’s d (pooled SD). Positive d indicates increase; negative d indicates decrease.*

**Table 3. Ceiling Effects (% at Maximum Score) by Scale and Timepoint**

| **Scale** | **Pre-op** | **1 week** | **3 months** | **1 year** |
| --- | --- | --- | --- | --- |
| Appearance Concerns | 16.1% | 16.4% | 26.3% | 31.3% |
| Scar Satisfaction | — | 19.2% | 31.3% | — |
| Psychosocial Distress | <5% | <5% | <5% | <5% |

*Values represent the percentage of respondents scoring at scale maximum. — = data not collected at this timepoint.*
